# Effectiveness of pain education on pain, disability, quality of life and self-efficacy in chronic low back pain: A randomized controlled trial

**DOI:** 10.1101/2023.10.31.23297833

**Authors:** Mohammad Sidiq, Tufail Muzaffar, Balamurugan Janakiraman, Shariq Masoodi, Rajkumar Krishnan Vasanthi, Arunachalam Ramachandran, Nitesh Bansal, Aksh Chahal, Faizan Zaffar Kashoo, Moattar Raza Rivzi, Ankita Sharma, Richa Hirendra Rai, Rituraj Verma, Monika Sharma, Sajjad Alam, Krishna Reddy Vajrala, Jyoti Sharma, Ramprasad Muthukrishnan

## Abstract

**Background:** Low back pain is one of the most common causes of pain-related disability worldwide. There are growing recommendations to use psychological approaches in the management of chronic low back pain. Pain education intervention is one such psychological approach aiming at re-conceptualizing pain beliefs and easing the pain threat value. This randomized controlled trial aimed to gain an understanding of the effectiveness of pain education on pain levels, disability, quality of life, and self-efficacy in individuals with chronic low back pain (CLBP).

**Methods:** A two-arm parallel randomized trial was conducted recruiting 92 participants with CLBP, who were randomly allocated to either standard physiotherapy care with the pain education program, or the control group, and both groups received 6 weeks of intervention. Pain intensity (using NPRS), disability (using RMDQ), self-efficacy (using general self-efficacy scale), and wellbeing (using WHO 5I) were assessed before, and 6 weeks after the study intervention.

**Findings:** The post-intervention scores comparison between the groups showed that the pain education intervention reduced disability compared to the usual standard care at 6 weeks (mean difference 8.2, p < 0.001, effect size η2 = 0.75), the pain intensity (mean difference 3.5, p < 0.001, effect size η2 = 0.82) and improved the wellbeing index (mean difference 13.7, p < 0.001, effect size η2 = 0.58).

**Conclusion:** The findings suggested that pain education program enhance the therapeutic benefits of usual standard physiotherapy care among participants with chronic LBP. We conclude that pain education seems to have clinical benefits when delivered along with standard care physiotherapy during the management of chronic low back pain.

**CTRI registration code:** CTRI/2021/08/035963

## Introduction

Chronic low back pain (CLBP) is a pervasive and debilitating condition affecting millions of individuals worldwide. It represents a significant global health burden, causing considerable pain, disability, and reduced quality of life for those affected. Chronic low back pain (CLBP) accounted for a total of 60.1 million person-years lived with disability [1,2]. Addressing the multifaceted nature of CLBP requires comprehensive treatment approaches that go beyond purely pharmacological interventions or a combination of pharmacological and physical rehabilitation [3,4]. Various non-surgical interventions have been provided to aid in the reduction of CLBP. These interventions include techniques like joint manipulation, acupuncture, traditional and contemporary therapeutic exercises, electrotherapy modalities, and medication [5].

However, these interventions have modest to no effect in addressing psychological barriers in recovery from low back pain [6]. The role of psychological factors in an individual’s experience of LBP is reported to have a bearing on their function, pain perception, belief in self-efficacy, and quality of life [3,6,7]. The current multimodal interventions in the management of CLBP recommend approaches involving pain education programs delivered with standard physiotherapy care [8,9]. Pain education is a psychological approach that focuses on improving the knowledge and understanding of pain using biological and neurophysiological-biomechanical explanations to impart reconceptualization of beliefs about the experience of pain particularly when it is chronic[10].

Although there is a growing recommendation to use pain education for chronic LBP, the challenge is that the pain education content exhibited variance in effectiveness based on the complexity of the curriculum, ethno-culture, individual pain experience context, and language needing examination [8,9,11]. By providing individuals with the knowledge and tools to better comprehend their pain experience, pain education interventions aim to reduce pain-related disability, improve quality of life, and increase self-efficacy in managing their condition. Since, the introduction of this form of pain education (‘Pain Neuroscience Education’ or ‘Explain Pain’) [12,13], it has been adopted by several Western countries with mixed findings for the efficacy of education. Further, literature [8,11,14,15] suggests that such psychological approaches that are effective in one culture may not be essentially effective in another. There is a scarce attempt of evaluation or adaptation of pain education material or implementation of pain education programs in the Eastern cultural context particularly in South Asia [15,16].

On the other hand, the effects of psychological approaches on chronic pain depend largely on the patients’ educational, ethnocultural, and social background which further limits the generalizability of the results of existing literature to different contexts, and warrants multi-cultural, contextual investigations in pain catastrophizing conditions like LBP. Therefore, this randomized controlled trial (RCT) seeks to determine the effects of pain education material on pain intensity, disability, quality of life, and self-efficacy among individuals with CLBP in India. The primary objective of this study is to evaluate whether pain education, delivered through a structured intervention, leads to a significant reduction in pain intensity compared to a control group receiving standard care. We also aim to investigate the secondary outcomes of disability, quality of life, and self-efficacy to assess the broader impact of pain education on individuals’ overall well-being.

## Methods

### Study design and Ethical consideration

This randomized controlled trial was conducted using a 2-arm parallel-group (1:1 allocation) design randomized controlled trial. This study used the Standard Protocol Items; Recommendations for Interventional Trials[17] statement during the protocol development (S1 file) and is reported in accordance with the CONSORT 2010 [18] guidelines (Figure 1). The protocol of the trial was approved by the Institutional Ethics Committee of SKIMS vide RP/114/2021 and was registered with the clinical trial registry of India (CTRI/2021/08/035963). All the methods of this trial were carried out in accordance with Declaration of Helsinki. All the participants were informed about the study purpose and aim, and were informed that the participation and withdrawal is voluntary. After the post-intervention outcome measure recording of the control group participants the pain education manual was provided to the control group with brief lecture for 10 minutes.

**Figure 1.**
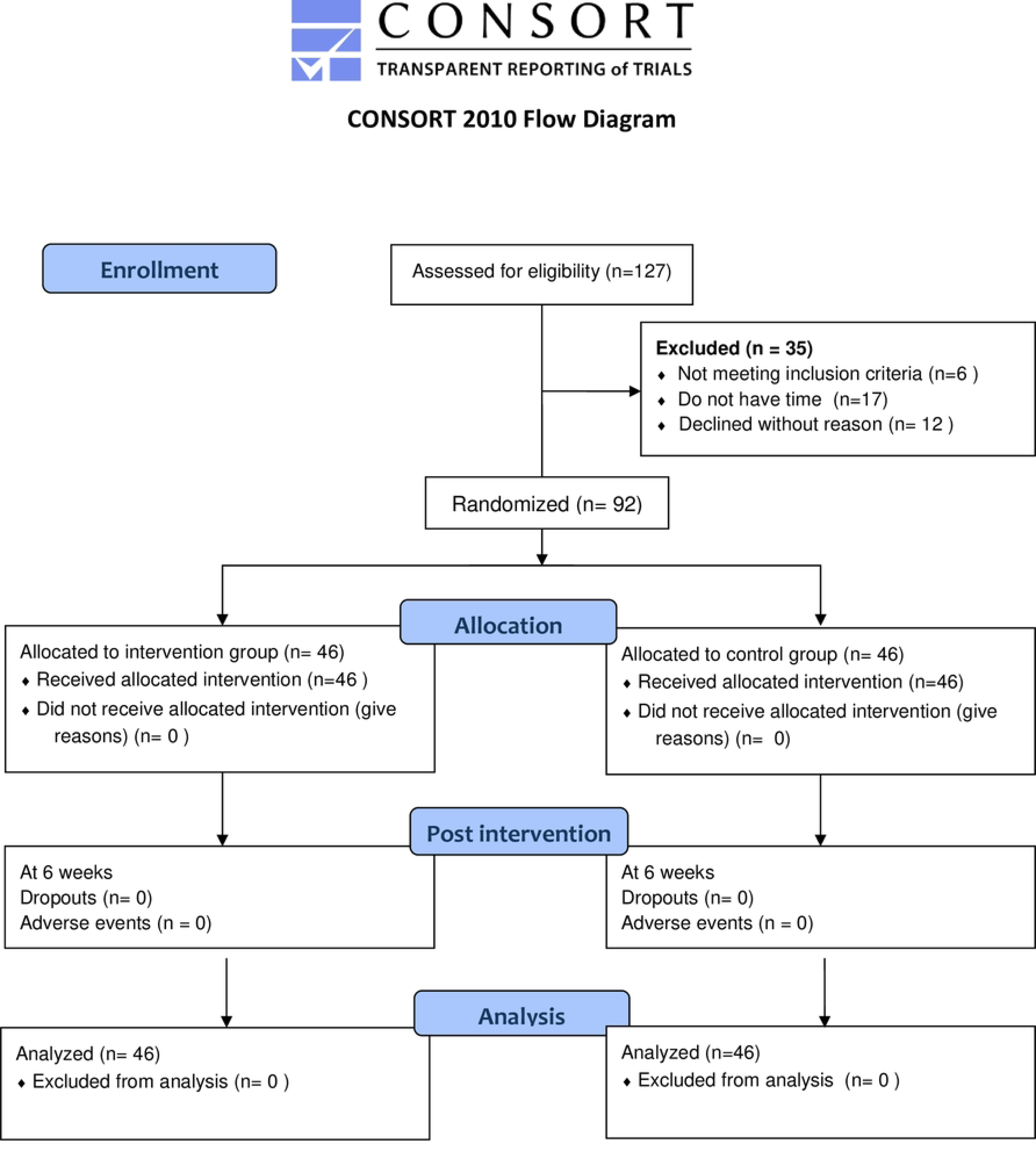
Consort participants flow diagram

### Participants and study setting

This study was conducted fromSeptember 2021 to October 2022 as a prospective parallel group active-controlled trial in the Department of Physical Medicine and Rehabilitation (PMR), Sher-i-Kashmir Institute of Medical Sciences (SKIMS), a tertiary public healthcare facility in Srinagar Kashmir India. The eligible participants were screened until the required sample of 92 patients were reached. The criteria for inclusion were: participants with nonspecific low back pain of more than 3 months duration diagnosed by the physiatrist based clinical examination and diagnostic procedures, aged between 18 to 60 years with both genders, and able to attend the study setting for all the intervention sessions, were included. The exclusion criteria were withdrawal from the study due to any reasons on voluntary basis, and not attending the intervention sessions more than twice The participants were blinded to their group assignment, as were the physiotherapists who completed the clinical outcome examinations. All participants were referred by the PMR physicians, they were informed about the vouluntary nature of the participation and informed consent was obtained from them. This trial adhered to the guidelines laid down by Helsinki Declaration [19].

### Sample size calculation

The required power calculated sample size was calculated using the following assumptions[20]; the confidence interval of 95%, a power of 0.80 (80%), and α error probability at 0.05 based on the findings of a similar study [21] regarding the mean score of pain intensity in chronic LBP patients (mean1 = 3.76, mean2 = 4.78, SD1 = 1.51, SD2 = + 1.91). Accordingly, the G power software version 3.1.9.4 for Windows the estimated sample was 46 participants per group.

### Randomization and blinding

A concealed allocation method using sequentially numbered, opaque, sealed envelops (SNOSE) [22] was used to randomly allocate the participants to the pain education group, or the standard physiotherapy care group. The office clerk of PMR department performed random allocation through block randomization with a block size of 4 and cards labeled A and B. The participants were blinded to the assignment of group, the assessors, and the data analyzer of the outcome measures were blinded about the intervention group assignment.

### Interventions

The participants in both the groups received routine standard physiotherapy care 6 weeks for the CLBP which consist of physical exercise and modalities. In addition, the pain education group received a fac-to-face education program consisted of modules defining chronic pain, neurophysiology of pain, and neurobiological aspects of pain including central sensitization, fear avoidance factors, and social factors affecting the experience of low back pain. The contents of the pain education was delivered by the first author to individual participants using powerpoint materials, images, lectures, and question and answer sessions for 2 session per week for the first 3 weeks, followed by question and answer sessions for the 4^th^ and 5^th^ week reflecting the learning, and at the end of the 6^th^ session of pain education, a brief pain education manual was provided to the participants of pain edication group. On average, the routine physiotherapy care lasted for 30 minutes for both the group.

#### Pain education manual development

A context and culture specific pain education manual was developed by the authors MS and BJ in Hindi language according the process recommended by Butler and Moseley [12,13,15]. The co-authors (SM, AC, FZK, NB) reviewed the Hindi pain education manual for the clarity and simplicity of the contents. The authors used literature related to pain education, clinical guidelines to treat LBP, pain stories in Hindi to explain target concepts of pain, and the pain education handbook (Explain Pain) was largely helpful in development of Hindi pain education manual. The final proof reading of the manual (S2 file) was conducted by a clinical psychologist, a physiatrist, a native Hindi speaking person, and a physiotherapist at SKIMS.

### Outcome measures

The data related to the socio-demographic characteristics of the study participants were recorded as per the recommendation of the national institute of health (NIH) task forceon research standards for chronic LBP [23]. The study outcomes were self-reported tools chosen to reflect the participants pain intensity, disability related to LBP, self-efficacy, and wellbeing.

#### Primary outcome measures

The primary outcome measures were related to the intensity of pain experienced during activities of daily life (ADL) and disability related to pain.Roland Morris Disability Questionnaire (RMDQ). The visual analogue scale (VAS) was used with the scores ranging from ‘0’ (no pain) to 10 (worst imaginable pain) on the scale and the participant was asked to mark on the scale. Which they flet represents the intensity of the pain while doing ADL. The 24-item Roland Morris Disability Questionnaire (RMDQ) was used to assess the disability, the RMDQ scores ranged from ‘0’ (no disability) to ‘24’ (high disability) [24].

#### Secondary outcome measures

The secondary study outcomes were those that potentially assesses the individual’s belief in their ability to cope with and succeed in challenging situations (General Self-efficacy Scale) and emotional wellbeing (World Health Organization Five Index) of the participants [25,26]. The General Self-efficacy Scale (GSE) is a self-reported measure of self-efficacy using 10-items, the scores range from ‘10’ to ‘40’ with higher scores indicating more self-efficacy. The GSE is a one dimensional outcome measure that assesses the optimistic self-beliefs of individual in coping with variety of demands in life and the World Health Organization-Five Well Being (WHO-5) is a self-reported 5- item measure of current mental well being. The WHO5 index raw score range from 0 to 25, with ‘0’ representing worst possible quality of life and ‘25’ representing best possible quality of life. The raw score is then multiplied by 4 to compute the percentage of WHO5 index percentage of quality of life (percentage score range 0 - 100). This trial expressed WHO 5 index as percent of quality of lfe.

## Data analysis

The patient characteristics at the baseline of this pain education intervention trial is presented as mean (SD), or frequency with percentage. The normality of the score distribution of all the outcome measures was tested with the Kolmogorov-Smirnov test and Levene’s was used to check the assumption of homogeneity. Unadjusted mean (SD), and standard error (SE) of the mean difference (MD) between the groups with associated 95% confidence interval and p-value were computed for the outcome measures at baseline and post-intervention. Literature reported potential prognostic variables like educational status, BMI, smoking, chronicity of symptoms (months), pain at baseline, disability (RMDQ) at baseline, WHO 5 Index, and GSE at baseline were included in the prognostic model. We performed a prognostic model logistic regression analysis by categorizing (dichotomous) the post-intervention scores at the closest value to the 75^th^ percentile (to represent 25% of those who did not improve versus 75% representing those who improved) [27]. The Post-intervention RMDQ score was dichotomized at ≥ 13, VAS at ≥ 5, WHO-5 well-being index at ≥ 60, and GSF at ≥ 33. These cutoffs used are designed to differentiate LBP patients who considered themselves not recovered versus those with high-level recovery and this method is recommended in literature in the absence of meaningful cut-off. The level of significance was set at 0.20 and 0.05 for univariate and multivariate logistic regression respectively to determine the main effect of potential prognostic variables (independent variables) on the outcome variable. The effect size of the intervention was calculated as the mean difference using Cohen’s d effect size. The level of significance was set at 5%. The IBM SPSS version 21 for Windows was used for analysis.

## Results

### Baseline characteristics

A total of ninety-two respondents diagnosed with chronic LBP by clinical examination or imaging, agreed to participate in this trial and until the completion of the intervention schedule period, none of them dropped out. Recruitment ran from 3^rd^ September 2021 to 9^th^ October 2022 prospectively. Table 1 shows the baseline characteristics of the experimental and control group participants. The mean age of patients in the experimental group was 42.0 years and that of the control group was 42.5 years. The variables exhibited statistical similarity between the experimental and control groups at baseline, except for the smoking habits and RMDQ scores. At baseline, the experimental group had a better mean score of RMDQ (RMDQ = 15.02, p 0.019, mean difference (MD) = 2.37), and also had more smokers than the control group (n = 11 (23.9%) versus n = 4 (8.7%), p 0.044). Participants of both the groups self-reported a statistically similar pain intensity of above 5 out of 10 in VAS at baseline (control group 5.98 and experimental group 5.7). The mean duration of chronicity of LBP among the participants was 7.4 and 7.8 months for the control and experimental group respectively. The secondary outcome measures WHO5 index and GSE were statistically similar between groups at baseline Table 1.

**Table 1.** Baseline characteristics on demographic factors, behavioural characteristics, clinical charaterisitics, and study outcome measures (n= 92)

| Variables | Control<br>($n = 46$) | Experimental<br>($n = 46$) | *P* |
| --- | --- | --- | --- |
| Age (years) | 42.5 ± 13.3 | 42.0 ± 8.2 | 0.829 |
| Gender (male/female)<sup>†</sup> | 11/35 | 20/26 | 0.077 |
| Occupation<sup>†</sup> |  |  | 0.531 |
| Self-employed | 14 (30.4) | 22 (47.8) |  |
| Housewife | 29 (63.0) | 15 (32.6) |  |
| Employee | 0 | 2 (4.3) |  |
| Student | 3 (6.6) | 7 (15.2) |  |
| Duration of LBP in months | 7.4 ± 1.2 | 7.8 ± 1.3 | 0.768 |
| Level of activity<sup>†</sup> |  |  | 0.681 |
| Very active | 9 (19.6) | 8 (17.4) |  |
| Moderate | 23 (50.0) | 20 (43.5) |  |
| Light | 14 (30.4) | 18 (39.1) |  |
| BMI (kg/m<sup>2</sup>) | 26.9 (4.0) | 27.8 (4.2) | 0.858 |
| Comorbidity<sup>†</sup> |  |  | 0.314 |
| None | 33 (71.7) | 37 (80.4) |  |
| Hypertension | 4 (8.7) | 5 (10.9) |  |
| Diabetes mellitus | 9 (19.6) | 4 (8.7) |  |
| Smoker<sup>†</sup> |  |  |  |
| Yes | 4 (8.7) | 11 (23.9) | 0.044 |
| No | 42 (91.3) | 35 (76.1) |  |
| Hours of sleep | 6.3 (1.7) | 6.7 (1.3) | 0.549 |
| Pain intensity (VAS 0-10) | 5.98±1.7 | 5.7±1.65 | 0.423 |
| Disability RMDQ ( 0-24) | 15.02±4.7 | 12.65±4.7 | 0.019 |
| WHO5I (0-100) | 54.4±18.6 | 53.5±17.5 | 0.804 |
| GSF (10-40) | 25.78±6.4 | 25.82±5.6 | 0.972 |

<sup>†</sup>Categorical variables are expressed as frequency and percentage, continuous data as mean and ± Standard deviation, <sup>†</sup> Chi-square test, LBP: low back pain, RMDQ: Roland-Morris Disability Questionnaire, WHO5I: WHO-5 Well-being Index, GSF: General Self-efficacy Scale

### Intervention findings

#### Primary outcomes

##### Disability

The patient education intervention using a structured booklet seemed to largely redu ce disability, measured by RMDQ, and intensity of pain, measured by VAS. The RMDQ score change within the group between baseline and post-intervention of both the groups were statistically significant (Experimental group; the mean difference (MD) 6.8, 95% confidence interval (CI) 0.4, 0.6, p < 0.01 and control group; MD 1.0, 95% CI 0.6, 1.3, p < 0.01) Figure 2.

**Figure 2.**
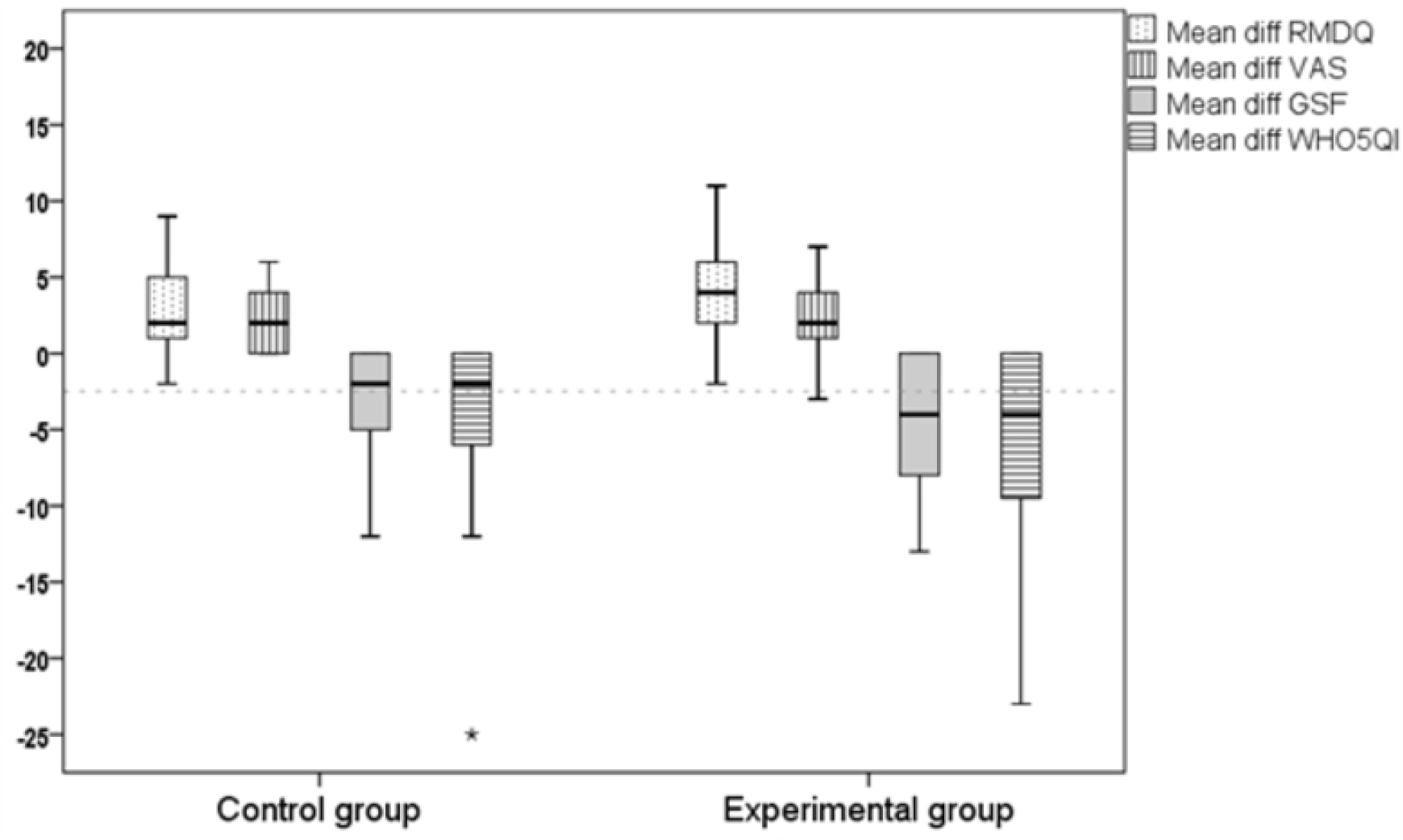
Mean difference of pre-test and post-test scores of RMDQ, VAS, GSF, and WHO5QI of the participants in the control group and experimental group.

The between-group mean difference in RMDQ score change at the post-intervention was 8.0, p < 0.001, with a large effect size (η2 = 0.75). The clinical change of RMDQ over time (6 months) was computed using ((baseline score minus post-intervention (MD) / baseline score) x 100). For the experimental group, at the start of the intervention, the RMDQ score was 12.65 and the immediate post-intervention score was 5.8. The calculated improvement of 6.85 points or 54.15% decrease in the level of disability. The mean RMDQ score of the participants in the control group was 15.02 at the start of the intervention and 14.0 immediately after the intervention period. The clinical change over time (6 weeks) was 1.02 points improvement or 6.8%. Pain education program led to a notable and statistically significant decline in disability related to LBP as reported by participants in the experimental group at 6 months post-intervention (Table 2).

**Table 02.** Unadjusted means, standard deviations, standard error, and 95% CI of the mean difference of the continuous outcome measures for control and experimental group (n =92)

| Outcome measures | Experimental<br>(n = 46) | Control<br>(n = 46) | p-value ($\eta^2$<br>value) |
| --- | --- | --- | --- |
| VAS |  |  |  |
| Post intervention | 2.1 (1.0) | 5.6 (1.5) | p <0.001 (0.82)<sup>a</sup> |
| Improvement (95%CI of MD) | 3.6 (3.2, 4.0) | 0.34 (0.2, 0.4) |  |
| SE of mean | 0.18 | 0.87 |  |
| Within-group difference p-value | p<0.01<sup>b</sup> | p<0.01<sup>b</sup> |  |
| RMDQ Score |  |  |  |
| Post intervention | 5.8 (3.4) | 14.0 (4.6) | p <0.001 (0.75)<sup>a</sup> |
| Improvement (95%CI of MD) | 6.8 (0.4, 0.6) | 1.0 (0.6, 1.3) |  |
| SE of mean | 0.40 | 0.15 |  |
| Within-group difference p-value | p < 0.01<sup>b</sup> | p < 0.01<sup>b</sup> |  |
| WHO-5 Score |  |  |  |
| Post intervention | 70.1 (14.7) | 56.4 (17.8) | p <0.001 (0.58)<sup>a</sup> |
| Improvement (95%CI of MD) | 17.1 (14.3, 20.0) | 2.0 (1.1, 2.8) |  |
| SE of mean | 1.41 | 0.41 |  |
| Within-group difference p-value | p < 0.01<sup>b</sup> | p < 0.01<sup>b</sup> |  |
| GSF |  |  |  |
| Post intervention | 28.8 (5.4) | 30.22 (4.36) | p 0.97 (-0.035)<sup>a</sup> |
| Improvement (95%CI of MD) | -3.04 (-4.11, -1.98) | -4.39 (-5.6, -3.09) |  |
| SE of mean | 0.52 | 0.64 |  |
| Within-group difference p-value | p < 0.01<sup>b</sup> | P<0.001<sup>b</sup> |  |

VAS-Visual analog scale; RMDQ-Roland Morris Disability Questionnaire; WHO-5-The World Health Organization-Five Well-Being Index, GSF: General Self-efficacy Scale, CI of MD- Confidence Interval of mean difference, <sup>a</sup> Between- group comparison of post intervention with analysis of covariance and effect size was assessed using the $\eta^2$ value, <sup>b</sup>within- group comparison before and after intervention with Paired Samples *t*-test.

**Table 03.** A backward stepwise logistic regression model with p values for each model variable predicting the main effects of prognostic variable (independent) on the outcome variable (post-intervention scores) at 6 weeks among the chronic LBP patients (n = 92) controlled for group allocation.

| Outcomes (at 6 weeks) | Prognostic predictors at baseline | $\beta$ | OR (95% CI) | $R^2$ | p-value for each model |
| --- | --- | --- | --- | --- | --- |
| Final disability (RMDQ≥ 13/24) | Education status | 0.689 | 1.99 (1.42, 3.86) | 0.21 | 0.008 |
|  | Disability | -0.29 | 0.75 (0.41, 0.89) |  | 0.012 |
| Final pain ($\geq 5/10$) | Pain intensity | 0.525 | 1.69 (1.31, 2.03) | 0.24 | 0.036 |
| Final WHO5 I ($\geq 60/100$) | Age | 0.79 | 2.20 (1.47, 3.81) | 0.19 | 0.002 |
| Final GSES ($\geq 33/40$) | Sex | 0.29 | 1.34 (1.09, 2.22) | 0.31 | 0.000 |
|  | Self-efficacy | 0.61 | 1.84 (1.46, 2.75) |  |  |

#### Pain

The intensity of pain was measured using the self-reported measurement tool (VAS), the scale can be completed in less than 1 min and the 10 cm horizontal line was labeled “no pain” on the left (assigned ‘0’), “worst imaginable excruciating pain” on the right (assigned ‘10’). The pre-/post-test scores of pain (VAS) between the experimental and control groups are shown in Table 2. Pain significantly alleviated within the experimental (mean difference 2.20. 95% CI; 1.55, 2.85, p < 0.01), and control groups (mean difference 2.435. 95% CI; 1.87, 2.99, p < 0.01), and a significant main effect with large effect size (p < 0.001, η2 = 0.82) between the two groups at post-intervention.

#### Secondary outcomes

WHO-5 well-being index differed significantly between baseline and 6 months intervention in favor of the pain education group with a moderate effect size (p < 0.001, η2 = 0.58). No significant difference was found for the GSE Table 2.

#### Regression model for prognostic predictors

Self-reported higher RMDQ at baseline increased the odds of elevated disability at 6 weeks (OR 1.99, 95% CI 1.42, 3.86), and higher educational status of the participants was protective against the higher disability at post-intervention (OR 0.75, 95% CI 0.41, 0.89). A higher VAS score at the baseline was associated with 1.69 folds (OR 1.69, 95% CI 1.31, 2.03) increased odds of higher pain intensity at 6 weeks. Better well-being and self-efficacy outcomes at 6 weeks were predicted by higher age (OR 2.20, 95% CI 1.47, 3.81), and female gender (OR 1.34, 95% CI 1.09. 2.22), higher self-efficacy at baseline (OR 1.84. 95% CI 1.46, 2.75) Table 03.

## Discussion

The pain education intervention to the chronic LBP patients has resulted in better outcomes in terms of pain, disability, and wellbeing among people with chronic LBP with effect sizes moderate to larger in 6 weeks term. The improvement of pain, wellbeing, and disability can be explained by the hypothesis that the pain education of LBP patients in the experimental group about realizing that most often pain is present without tissue damage due to central sensitization and thinking more about movement than pain [28,29]. This would have probably led to a shift of patients opinion about LBP and develop a self-management bio-psychosocial approach which is claimed to reduce pain LBP by decreasing central sensitivity [30,31]. Additionally, higher self-reported disability and high pain intensity were found to be indicators of poor prognosis in this trial [32,33].

Participants with higher educational attainment and females had higher probability reporting better wellbeing and self-efficacy at 6 weeks of intervention. This is rather not surprising, because higher education would give more insight and better understanding into the pain education program, helping them to cope pain, and annihilate the challenges related to LBP [34,35]. However, overall there was a significant improvement in the primary outcomes (RMDQ and VAS) and wellbeing in the pain intervention group compared to the control group. Surprisingly, no meaningful association was found in logistic regression model between the prognosis of outcome measure scores and smoking habits, whereas few literatures [27,32,36] identified smoking as a prognostic indicator in LBP patients. Similar to the findings of this study, a cognition targeted intervention study [37] reported improvement of pain and disability in individuals with spinal pain. Their education based intervention demonstrated larger ES (0.66 and 0.81 for pain and disability respectively. Likewise, in this study the results after 6 weeks intervention showed ES of 0.82 for pain, and 0.75 for disability.

The ES for pain and disability followed by pain education intervention varied across trials, Malfliet et al [14] reported a lower ES of 0.52 for pain and 0.49 disability at 3 month follow-up compared to this trial. In contrast, a pilot trial by Ibrahim et al [38] reported a larger ES for disability (2.22) and pain (1.66) employing group patient education combined with motor control exercise as intervention to low resource rural community dwelling adults with chronic LBP. The clinical change in RMDQ scores demonstrated a substantial improvement in our experimental group, with a 54.15% decrease in disability levels compared to a 6.8% improvement in the control group. Notably, an imptovement of 30% and more in RMDQ score is rated as clinically relevant [39]. This findings further strengthens the claim on the effectiveness of the pain education intervention in reducing disability related to CLBP which is similar to studies reporting efficacy of pain education interventions [10,21,35,37]. Regarding pain intensity, as measured by the Visual Analog Scale (VAS), both the experimental and control groups showed a significant decrease following the intervention. However, the mean difference in pain intensity score change between the groups remained statistically significant after adjusting for confounding variables, with a larger effect size observed in the experimental group. This indicates that the pain education program led to a notable and statistically significant decline in pain intensity reported by participants in the experimental group at the 6-month post-intervention assessment.

Furthermore, the pain education intervention had a positive impact on the well-being of participants, as indicated by the WHO-5 well-being index. The experimental group in this trial demonstrated a significant improvement in well-being from baseline to the 6-month intervention period, with a moderate effect size. This positive effect on well-being remained consistent even after accounting for potential confounders and evidence suggest that psychological interventions being effective in improving wellbeing [40]. However, in this study there were no significant differences between the experimental and control groups in terms of general self-efficacy. Further, studies report association of low self-efficacy among adults with CLBP and the need for interventions [41].

Pain education programs delivered along with conventional treatment has shown significant improvement in pain reduction[10,14] as compared to the other group. Similarly, two reviews [10,42] reported that pain education enhances pain reconceptualization, which might facilitates patient’s ability to cope with their conditions. However, one of those review[42] found that pain education has only short-term effects when used long side physiotherapy interventions in CLBP. Unfortunately, this study did not record the outcome measures at follow up to investigate the retention effects of pain education intervention. The group with pain education has shown significant improvement showing that the comprehensive approach targeting both physical and psychological aspects of pain management offers promise for improving disability in individuals with chronic low back pain. Simiral findings were reported by the studies conducted by Saracoglu et al, 2020 and Malfliet, A et al, 2017 [14,43] addressing that psychological and neurophysiological aspects of pain education enhances the understanding and management of pain, resulting in reduced pain levels, improved physical function, and decreased disability.

The pain education is reported to be effective in reducing self-reported disability and pain in chronic conditions like cancer, osteoarthritis, post-operative, and associated symptoms based on the findings of previous literature[44,45]. The findings of this study also demonstrated that the partcipants quality of life and wellbeing improved with pain education. A better quality of life and an improved ability to engage in daily activities and pursue meaningful goals in need of patients with chronic low back pain. The importance of educating patients about the underlying mechanisms of pain to improve their pain management and overall quality of life by pain education was emphasized by Louw and others [46]. The study by Mosely et al. 2014, highlighted the effectiveness of intensive neurophysiology education in reducing pain intensity and improving physical functioning and psychological well-being in individuals with chronic low back pain thus promoting overall well-being [47]. Malfliet el al, also reported that quality of life can be improved by delivering pain education with motor control training in patients with low back pain [14]. Similarly, a cross-sectional study, reported that pain self-efficacy accounts for a greater degree of variation in disability compared to fear of movement. Further, it was observed that changes in self-efficacy, rather than changes in fear of movement, serve as a mediator between changes in pain intensity and changes in disability over the course of one year [48]. More importantly, pain education intervention can be tailored for individuals or to suit a particular group of individuals experiencing chronic pain. Interventions by pain education could be delivered outside the healthcare system at a very low resource set up, these characteristics of pain education suggest that it can be effective, versatile, and alternative to current practice.

### Strength and limitations

To the best of our knowledge, this trial is one of the fewest adequately powered study in India to have prospectively registered, with design featured to minimize possible bias and provide insight into the possible changes the pain education has on the pain, disability, and wellbeing among chronic LBP patients. Further, this study had a control group treated with a standardized plan of care in contrast to a few trials which offered minimal or usual care control interventions. There are a few limitations worthy of discussion and will help execute caution while interpreting the findings of this study. First, the linguistically and culturally diverse population in India may demand tailored pain education program specific to the population, emphasizing diverse needs for most behavioral treatment options. Further, the feasibility constraints did not allow follow up data recording, and hence the retention effect of the pain education ptogram in this study is not known.

## Conclusions

LBP has always been a significant source of disability and develops into chronic pain more often than not. Pain education seems to be a favorable intervention in alleviation of pain and reducing disability in chronic LBP. Yet future studies are warranted to conclude the efficacy of pain education among different socio-economic, ethno-culturally diversified strata of individuals, people with different educational attainment, and even examine the feasibility of online or tele delivery of pain education.

## Data Availability

All the data supporting the findings of this trial is presented and the full dataset are available from the principal investigator on request.

## Declaration

### Author contributions

MS conceptualized, designed, and developed the pain education manual package. MS, SM and TM were involved in traning the data collectors, BJ and MS conducted data curation, the analyses, and interpretation of data, MS, BJ, and SM drafted the article. All the authors critically revised the article and provided important intellectual content. All the authors discussed the results, and approved the final version of the manuscript.

## Acknowledgment

Authors would like to thank the Sher-i-Kashmir Institute of Medical Sciences (SKIMS) for the permission and support to conduct this study. We are thankful to the physiotherapists involved in the interventions and the assessors for helping in data collection. Finally, we express out gratitude to the researchers of SKIMS for their valuable suggestions, and all the technical staffs associated with this study.

